# Widespread testing, case isolation and contact tracing may allow safe school reopening with continued moderate physical distancing: a modeling analysis of King County, WA data

**DOI:** 10.1101/2020.08.14.20174649

**Authors:** Chloe Bracis, Eileen Burns, Mia Moore, David Swan, Daniel B Reeves, Joshua T. Schiffer, Dobromir Dimitrov

## Abstract

**Background:** In late March 2020, a “Stay Home, Stay Healthy” order was issued in Washington State in response to the COVID-19 pandemic. On May 1, a 4-phase reopening plan began. If implemented without interruptions, all types of public interactions were planned to resume by July 15. We investigated whether adjunctive prevention strategies would allow less restrictive physical distancing to avoid second epidemic waves and secure safe school reopening.

**Methods:** We developed a mathematical model, stratifying the population by age (0-19 years, 20-49 years, 50-69 years, and 70+ years), infection status (susceptible, exposed, asymptomatic, pre-symptomatic, symptomatic, recovered) and treatment status (undiagnosed, diagnosed, hospitalized) to project SARS-CoV-2 transmission during and after the reopening period. The model was parameterized with demographic and contact data from King County, WA and calibrated to confirmed cases, deaths (overall and by age) and epidemic peak timing. Adjunctive prevention interventions were simulated assuming different levels of pre-COVID physical interactions (pC_PI) restored. We made several predictions related to adjunctive interventions or changes in pC_PI.

**Results:** The best model fit estimated ~35% pC_PI under lockdown. Gradually restoring 75% pC_PI for all age groups between May 15-July 15 resulted in ~350 daily deaths by early September 2020. Maintaining less than 45% pC_PI was required with current testing practices to ensure low levels of daily infections and deaths. If widespread community transmission persisted, isolating the elderly does not lower daily death rates significantly. Increased testing, isolation of symptomatic infections, and contact tracing permitted 60% pC_PI without significant increases in daily deaths before September, although this strategy may not be sufficient to eliminate community transmission. This combination strategy also allowed opening of schools with <15 daily deaths. Inpatient antiviral treatment reduces deaths significantly without lowering cases or hospitalizations.

**Conclusions:** We predict that widespread implementation of “test and isolate” policy alone is insufficient to prevent the rapid re-emergence of SARS CoV-2 without moderate physical distancing. However, widespread testing, contact tracing and case isolation would allow relaxation of physical distancing, as well as opening of schools, without a surge in local cases and deaths.

## Introduction

The current COVID-19 pandemic started in Wuhan China in late December 2019. It rapidly spread across the globe soon thereafter and has disrupted normal life in virtually every country in the world ever since. As of July 30, more than 17 million confirmed cases had been reported around the world resulting in close to 670, 000 deaths.^1^ While the scientific community has been is focused its full attention on developing effective treatments and vaccines, physical distancing – in many cases including quarantine of suspected and confirmed cases and contact tracing – has been the only effective prevention approach to reduce local attack rates.

In May, in the United States (US), many local and national governments developed plans to relax lockdowns and restore the sense of normalcy in their communities. These plans sought to delicately public health with economic and societal health. Vital societal institutions like workplaces and schools, were deemed worthy of reopening with measures designed to prevent rapid resurgence of infections and deaths. Unfortunately, results from many states where cases are mounting suggest that this process was too abrupt.^2^ However, other countries in Europe and Asia, as well as the Northeastern United States, have demonstrated the possibility of careful, safe reopening, including schools, bars, and restaurants, without a massive surge in cases.^2,3^

Washington State (WA) holds a special place in the history of the COVID epidemic with both the first US case of COVID-19 (Jan 20) and the first death due to COVID-19 (Feb 29).^4^ Shortly thereafter, state authorities began imposing travel and gathering restrictions and many local businesses started implementing “work from home” policies. The process culminated with the “Stay Home, Stay Healthy” order of the Governor issued on March 23.^5^ On May 1, a plan for reopening in 4 phases was announced which, if implemented without interruptions would have resumed all public interactions with physical distancing by July 15.^6^ This plan was consequently updated multiple times with the majority of WA counties (including King County, home of the Seattle metro area) have not progressed beyond phase 2 as of July 30 due to ongoing widespread incident infection.^7^

Mathematical models have been employed to project the course of COVID-19 outbreaks in different settings and to inform policymaking at the local and national level.^8-13^ However, models require specific parameterization, depending on the geographic and political context. This procedure unavoidably makes critical assumptions that cannot be informed by local data. For instance, wide parameter ranges associated with asymptomatic infections (both prevalence and infectiousness) help fit models to cumulative case and death counts. However, varying these parameter values leads to significantly different estimates of the seroprevalence in the population at the end of the outbreak (final size), an essential prediction for those planning reopening strategies. The large uncertainties related to asymptomatic infections is recognized by CDC and found its way into the COVID-19 pandemic scenarios designed to help inform ongoing modeling studies.^14^

While the majority of early studies focused on estimating the effects of physical distancing and projecting the burden on healthcare systems during the initial outbreak, the focus has switched to evaluation of potential reopening scenarios in different settings. In this study, we use a mathematical model, specifically calibrated to King County, to project SARS-CoV-2 transmission during and after various reopening scenarios. We quantitatively investigate adjunctive interventions such as early test and isolate, early test and treat, and post exposure prophylaxis. Our primary goal was to understand how adjunctive interventions and physical distancing, may be used to help society return to “normalcy” and endure potential subsequent epidemic waves. We also address the critical question of school reopening in fall 2020 and its potential impact on the epidemic situation in King County.

## Methods

### Model description

We use a deterministic compartment model to describe epidemic dynamics. Our model (**Fig 1 and Fig S1**) stratifies the population by age (0-19 years, 20-49 years, 50-69 years, and 70+ years), infection status (susceptible, exposed, asymptomatic, pre-symptomatic, symptomatic, recovered) and treatment status (undiagnosed, diagnosed, hospitalized). The forces of infection, representing the risk of the susceptible individuals by age to acquire infection (transition from susceptible to exposed), are differentiated by age of the susceptible individual, the contact matrix (proportion of contacts with each age group), infection and treatment status (asymptomatic, pre-symptomatic, symptomatic, diagnosed and hospitalized cases) of the infected contacts, and the time-dependent reduction of transmission due to physical distancing measures (work from home, closing non-essential businesses, banning large gathering, etc,) applied in the area (scaled up starting March 8 and fully taking effect March 29) and later relaxed during the reopening after May 15. A full description of the model can be found in the Supplement.

**Figure 1.**
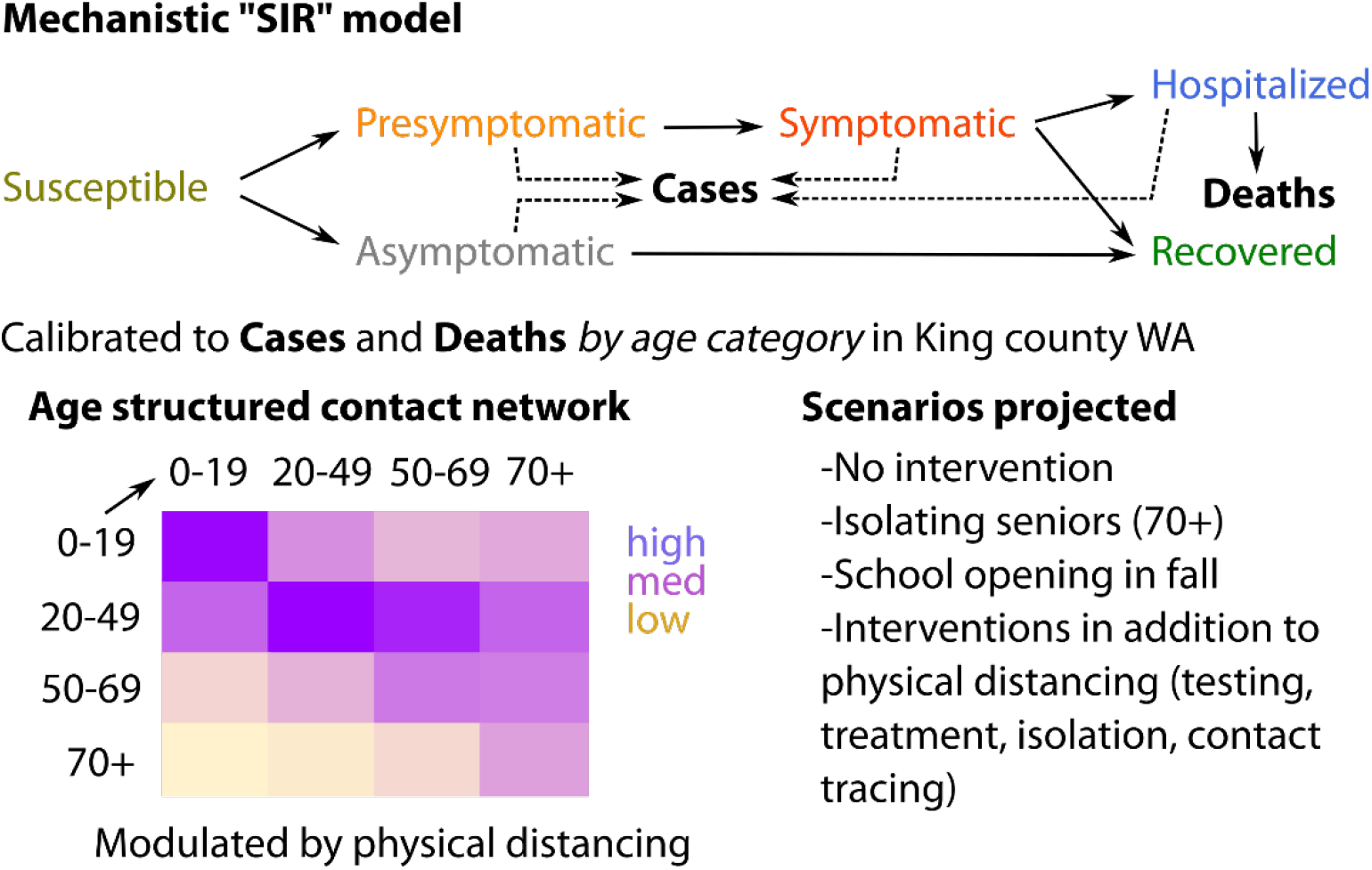
Simplified diagram of the modeling analysis.

### Parameterization, calibration and validation

The model is currently parameterized with local demographic and contact data from King County, WA and calibrated using transmission parameters priors informed from published sources. The model is calibrated to 5 “targets” based on local data (**Fig 2**), including: i) overall number of confirmed cases (target #1) and deaths (target #2) reported in King County over time since the start of the epidemic outbreak through April 30; ii) age-distribution of the cumulative confirmed cases (target #3) and deaths (target #4) reported in King County at 3 time points after the start of the epidemic outbreak and iii) the timing of the peak of daily confirmed cases (target #5) estimated as April 1.^15^ We used a genetic algorithm (NSGA-II multivariate optimization algorithm in the mco R package) to evolve a population of parameterizations to arrive at a set approximating the Pareto front. We defined thresholds for each target, based on a reasonable fit to that target and used them to select the best fit from the final population.

**Figure 2.**
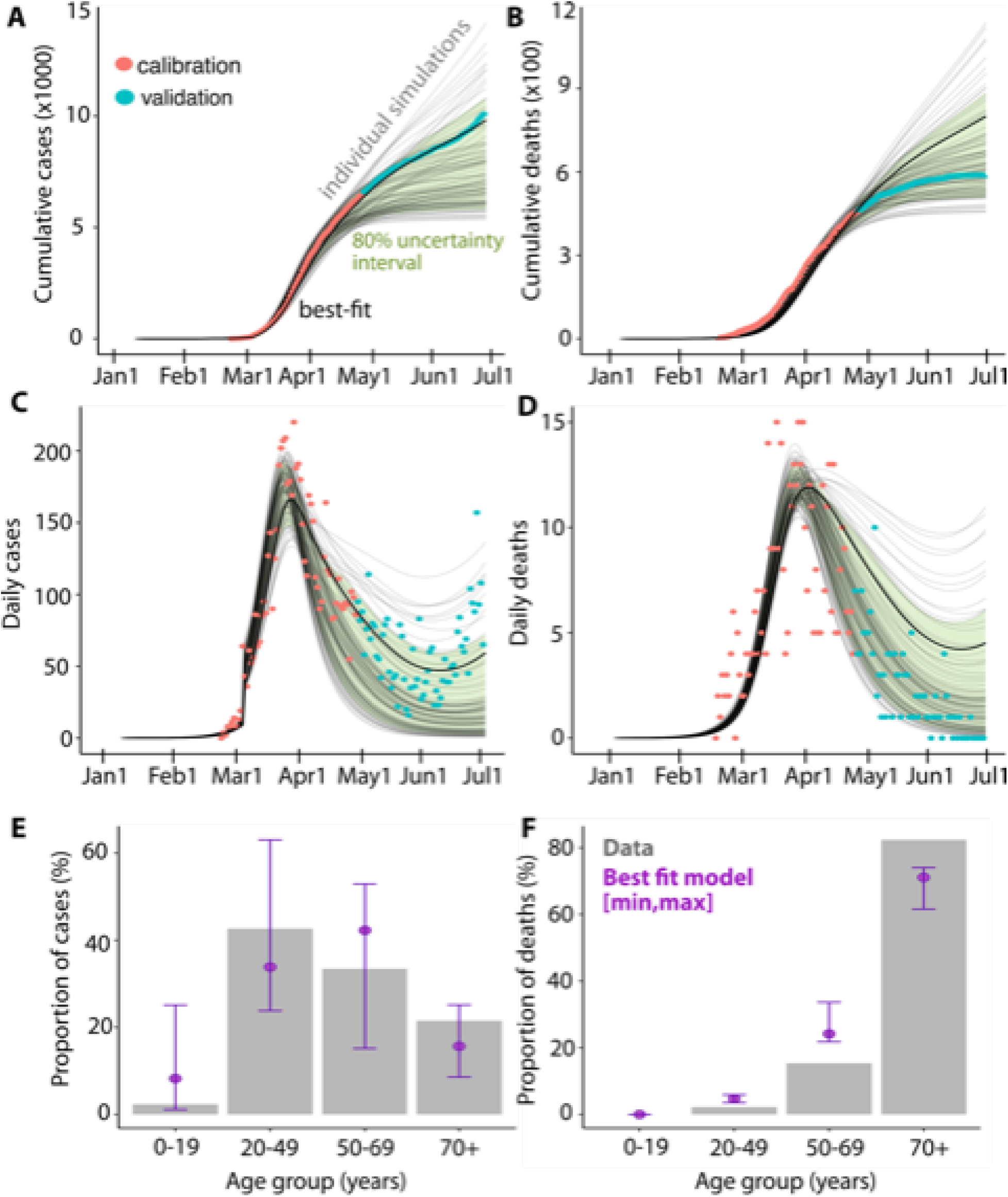
Model calibration and validation. Model fitting to 5 sources of King County data assuming gradual scale up of social distancing between March 8 and March 29: A)-B) Cumulative and daily cases and deaths. Red dots represent data up to April 30, thick lines represent the best model fit while other acceptable trajectories are shown in grey. Green bands show 80% range from acceptable trajectories. C)-D) Age distributions of cases and deaths as of April 15. Bars represent data, green dots and ranges represent the best fit and other acceptable trajectories included in the analysis. Reopening plan is implemented between May 15 and July 15 by <u>restoring 60% of pC PI</u> in all age groups.

Next, we used Monte Carlo filtering to select 100 parameter sets which reproduce data within these pre-specified tolerances. Resulting sets were used to explore the uncertainty in our model projections (see **Fig S2**). More details on the calibration scheme are in the Supplement.

We validate our population model by predicting independent data not used for calibration: i) cumulative number of confirmed cases and deaths between April 30 and June 30, as well as estimated number of daily hospitalizations and the overall hospitalization rates among confirmed cases at the end of April; ii) expert predictions informed by seroprevalence data of the cumulative incidence. We also compare to independent region-specific modeling projections including cumulative SARS-CoV-2 incidence at the beginning of March based on genomic analyses^16^ as well as the cumulative incidence and effective reproductive number (Rt) estimates for King County for the period between March 1 and April 22 reported by the Institute for Disease Modeling^17^.

### Reopening plans and intervention scenarios

Given the impact of the COVID epidemic on daily life and recommendations from the Centers of Disease Control (CDC) aiming to prevent new infections with continued physical distancing, hand washing, and masking, it is reasonable to expect that even after a fully implemented reopening plan, transmission reduction measures will remain in place. We thus assume reduction in transmission of at least 25% in comparison to early (pre-lockdown) levels. This scenario represents a reasonable limit because our projections suggest an unreasonably severe epidemic at this level of physical distancing. We explore reopening scenarios assuming gradually restoration of up to 75% of the pre-COVID physical interactions (pC_PI) for all age groups between May 15-July 15 (see **Fig 3**) with baseline scenario assuming 60% pC_PI. We also simulate an alternative scenario (Protect seniors) in which the oldest age group (70+ year-old) remain under strict physical distancing with pC-PI at lockdown levels. Finally, we evaluate the impact of school reopening on September 1 under baseline or “Protect seniors” reopening plans.

**Figure 3.**
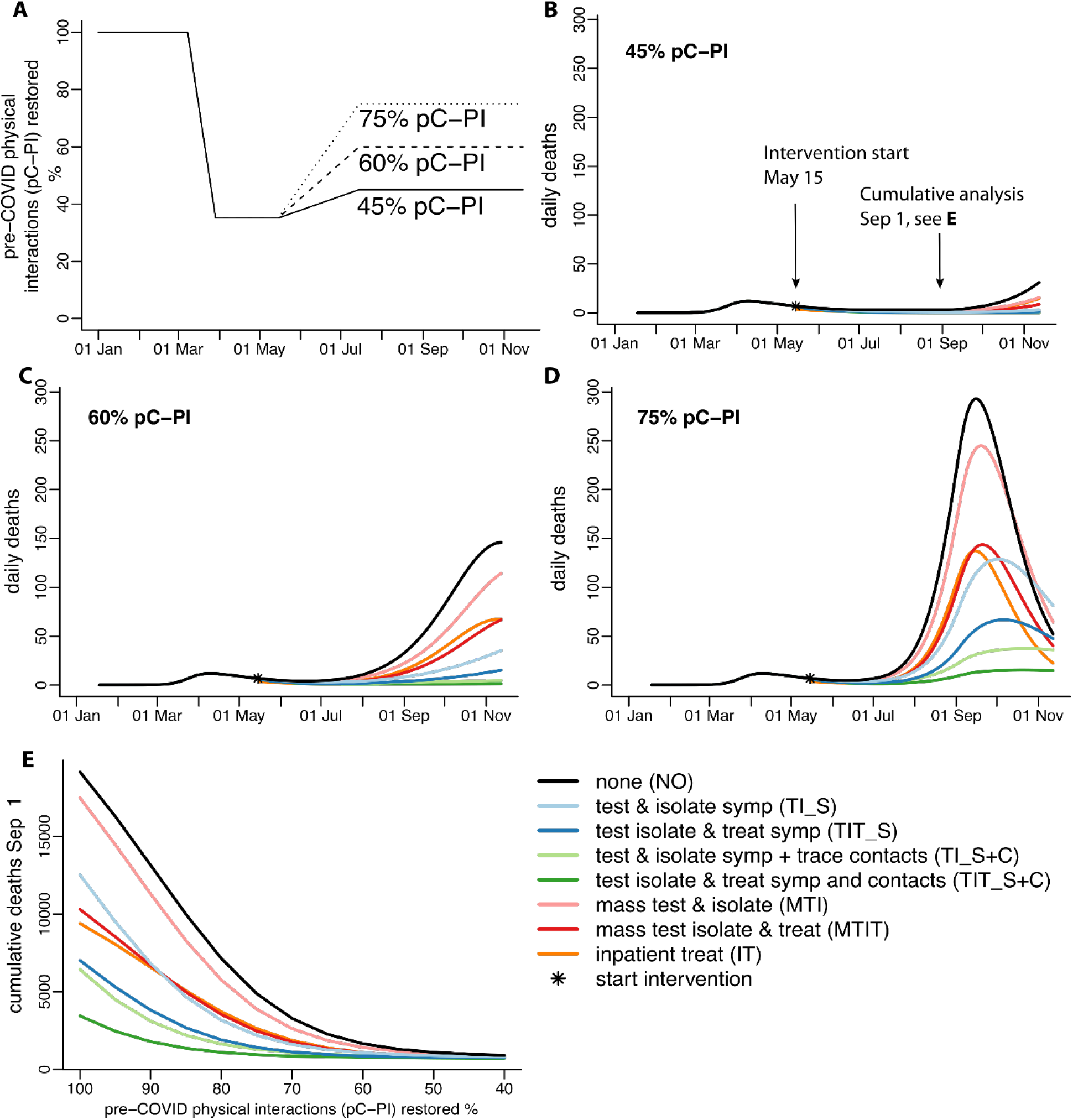
Model best fit projections under different intervention scenarios. **A**) Time variation of the physical interactions for specific levels of pC_PI restored B-D) Daily deaths for specific levels of pC_PI restored; E) Cumulative deaths from the beginning of the outbreak to Sep 1. Reopening plan is implemented from May 15 to July 15 by restoring different levels of pC_PI as shown in the upper left corner of each panel.

Several adjunctive prevention interventions listed in **Table 1** were simulated assuming different levels of pre-COVID physical interactions (pC_PI) restored during reopening. These include non-pharmaceutical interventions which are currently available such as rapid and mass testing, isolation and contact tracing; as well as hypothetical treatment options for exposed individuals (post-exposure prophylaxis), mild cases (outpatient treatment) and severe cases (inpatient treatment). In short, we assume that: i) enhanced early testing will increase the daily probability that symptomatic individuals get tested to 10% in the main scenario which will ensure that more than 50% of the symptomatic infections (more than 40% of all infections) are diagnosed. Estimates based on serological data from late March suggest that less than 10% of all infections are reported.^18^ Even more effective programs of early testing assuming symptomatic diagnostic rates up to 50% daily are also evaluated; ii) contact tracing allows for testing 5% of the asymptomatic and pre-symptomatic cases in the main scenario assuming that 50% of the contacts of the diagnosed cases will be traced; iii) random mass testing will add 0.5 percentage points to the diagnostic rates among asymptomatic, pre-symptomatic and symptomatic cases which implies that at least 10,000 random tests are performed daily. Mass testing of up to 4.5% of the total population was explored which at the upper bound would mean that the entire population is tested monthly. We also assume that imperfect isolation will halve the transmission from diagnosed cases from its lowest level during lockdown while effective early treatment will halve it again and reduce hospitalization rates among diagnosed by 50%. Finally, effective inpatient treatment is assumed to improve the recovery rate by 20% and reduce mortality rates by 50% similar to reported data from a clinical trial of remdesivir.^19^ Full descriptions of the reopening and intervention scenarios are provided in Table 1.

**Table 1.** Reopening and intervention scenarios.

| Scenario (NAME) | Simulated effects in the model |
| --- | --- |
| **<u>Reopening plans:</u>** |  |
| No intervention, uniform expansion of physical contacts across age groups (**Baseline**) | Gradually increase physical interactions to predefined post-reopening levels over 2-month period (May 15-July 15) <u>for all age groups</u> assuming that diagnostic rates remain unchanged. |
| Extended physical distancing for seniors (**Protect seniors**) | Gradually increase physical interactions to predefined post-reopening levels over 2-month period (May 15-July 15) for age groups 1,2 and 3 only. All diagnostic rates remain unchanged. |
| Schools reopening in fall (**Reopen schools**) | Additional increase of the physical interactions of the youngest group to 80% of the pre-COVID on Sept.1 |
| **<u>Interventions:</u>** |  |
| Effective inpatient treatment (IT) | Reduce mortality rate among hospitalized by 50%<br>Improve recovery rate among hospitalized by 20% |
| Rapid test and isolate symptomatic (**TI_S**) | Increase diagnostic rates among symptomatic to 10% daily (Rates up to 50% explored)<br>Reduce transmission from diagnosed by 50% |
| Rapid test, isolate and treat symptomatic (**TIT_S**) | Increase diagnostic rates among symptomatic to 10% daily (Rates up to 50% explored)<br>Reduce transmission from diagnosed by 75%<br>Reduce hospitalization rate from diagnosed by 50% |
| Rapid test and isolate symptomatic + trace, test and isolate contacts (**TI_S+C**) | Increase diagnostic rates among symptomatic to 10% daily (Rates up to 50% explored)<br>Reduce transmission from diagnosed by 50%<br>Increases diagnostic rates among asymptomatic and pre-symptomatic to 5% daily |
| Rapid test, isolate and treat symptomatic + trace, test and treat contacts (**TIT_S+C**) | Increase diagnostic rates among symptomatic to 10% daily (Rates up to 50% explored)<br>Reduce transmission from diagnosed by 75%<br>Reduce hospitalization rate from diagnosed by 50%<br>Increases diagnostic rates among asymptomatic and pre-symptomatic to 5% daily |
| Mass testing and isolate (**MTI**) | Increase diagnostic rates among asymptomatic, pre-symptomatic and symptomatic by 0.5 percentage points daily (Increase up to 4.5 percentage points explored)<br>Reduce transmission from diagnosed by 50% |
| Mass testing, isolate and treat (**MTIT**) | Increase diagnostic rates among asymptomatic, pre-symptomatic and symptomatic by 0.5% percentage points daily (Increase up to 4.5 percentage points explored)<br>Reduce transmission from diagnosed by 75%<br>Reduce hospitalization rate from diagnosed by 50% |

### Metrics of interest

We evaluate the course of the epidemic and effectiveness of different adjunctive interventions by tracking several key metrics including cumulative and daily number of deaths, number of new and current hospitalizations and effective reproductive number (Rt). Point estimates based on the “best fit” are presented in addition to 80% uncertainty interval (UI) based on 100 acceptable trajectories simulated per scenario. We report the results annotated by BF for the ‘best fit’ parameterization and by UI when referring to the sampled set of 100 parameterizations.

### Sensitivity analysis

Some of the key unresolved uncertainties related to SARS-CoV-2 transmission concern the prevalence and infectiousness of the asymptomatic infections. Our literature review has shown that proportion of infections which progress without symptoms is estimated from 10% to 70% by different empirical and modelling studies.^20,21^ Recently, asymptomatic transmission was qualified as “very rare” by one WHO official who later clarified that “the actual rates of asymptomatic transmission aren’t yet known.^22^ Previous studies demonstrated that asymptomatic infections have periods of viral shedding allowing for transmission. However, these individuals often have lower viral loads, so it is unclear if transmissibility is identical to symptomatic shedders.^23,24^ More importantly, very few infected people, symptomatic or asymptomatic, have been sampled during the earliest phase of infection when contagiousness is likely to be highest. In our main scenario we assume that 20% of the infections are asymptomatic and that they are as infectious as the symptomatic infections (100% relative infectivity). In sensitivity analysis we vary the prevalence of asymptomatic infections between 10% and 50% and relative infectivity of asymptomatic infections between 50% and 100% in unison with the CDC recommendations.^14^

## Results

### Effectiveness of physical distancing during the early King County outbreak

We generated an ensemble of 100 model parameterizations that recapitulated cumulative and incident deaths and cases stratified by age cohort (**Fig 2**). Projected peak of case numbers and hospitalization precede numbers of deaths by approximately 9 days. Our best fit (BF) suggests that physical distancing reduced SARS-CoV-2 transmission by 65% when fully implemented in King County with a range of 54%-83% (UI) from the sampled parameter sets. We estimated that the effective reproductive number (Rt) decreased from 2.43 (BF, range 2.19-2.55 UI) at the start of the epidemic to 0.81 (BF, range 0.40-0.93 UI) by the end of April and remained below 1 at the time when reopening started for all parameterizations.

Had transmission not been reduced by physical distancing, we estimate that by May 15, more than three quarters of King County residents would have been infected with 17300 additional excess deaths (BF). Our model suggests that the virus was likely introduced into King County around Jan 15 and projects approximately 2% (BF, range 1-2% UI) cumulative incidence among the population in King County by May 15 including those infected and recovered. We estimate that only 21% (BF, range 18-34% UI) of the symptomatic infections through May 15 were diagnosed.

### Projections of reopening plans with no added interventions

Degree of restoration of pre-contact physical interactions (**Fig 3A**) is the key determinant of severity of local outbreaks in the absence of adjunctive interventions. Our model suggests that daily deaths and cases would remain low without additional interventions if physical interaction is kept at 45% of the pre-COVID level even without additional interventions (**Fig 3B**).

With gradual progression towards 60% pC-PI, cases are predicted to surge starting in mid-July, which is consistent with current observations (**Fig 2B**). At 60% pC-PI, the cumulative number of deaths in the absence of intervention is expected to be ~2000 by November 1 (**Fig 4A**) with a peak of deaths per day far exceeding the peak of 15 deaths per day observed in April (**Fig 3C, 4B**). In half of the simulations, the number of current COVID-19 hospitalizations is expected to surpass 1700 (**Table S5**) exceeding more than three times the state mandated maximum of 10% occupancy. We predict that that the threshold of 15 deaths per day would likely be met in October but possibly as early as August under these conditions (**Fig 4C, black**).

**Figure 4.**
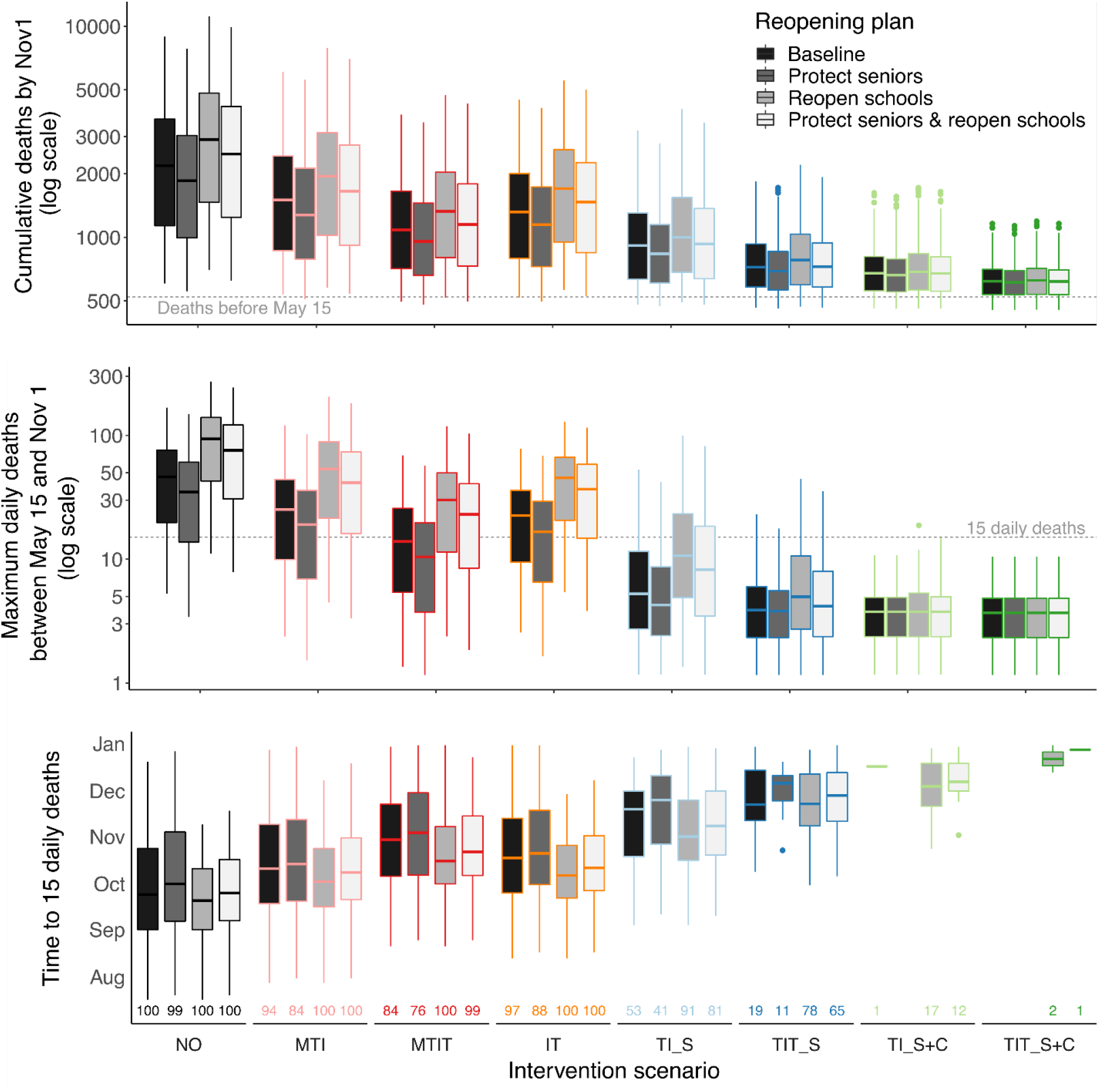
Model projections under different combinations of reopening scenarios and adjunctive interventions in terms of. A) Cumulative deaths by Nov 1. B) Maximum daily deaths by Nov 1; C) Time from the start of reopening to reach 15 deaths daily. Reopening plan is implemented between May 15 and July 15 by <u>restoring 60% of pC PI</u> in all age groups (Baseline) or age groups 1-3 only (Protect seniors). Schools reopen on Sept.1.

A peak of ~300 daily deaths per day in September is projected for 75% pC_PI (**Fig 3D**). pC_PI exceeding 55% is predicted to be associated with a high number of daily deaths by September 1 in the absence of an additional intervention (**Fig 3D**).

### Predicted failure of all adjunctive interventions with restoration of 75% pre-contact physical interactions

When applied at 75% pC-PI, test and isolate strategy (TI_S) is unlikely to prevent a massive epidemic resurgence with nearly 400,000 diagnosed cases and more than 15,000 deaths by November 1 with a slightly delayed peak of ~150 daily deaths in early October (BF, **Fig 3D** and **Fig. S4**, light blue). Therefore, resumption of lockdown levels of physical distancing would have been necessary in mid to late July under these conditions. With restoration of 75% of pC-PI, only the implementation of the most comprehensive strategy (TIT_S+C) including early testing, contact tracing, isolation and COVID-19 treatment of cases and contacts (**Fig 3D, dark green**) may prevent unacceptable levels of excess deaths with notable improvements relative to testing and treatment of symptomatic cases (TIT_S) and testing and tracing (TI_S+C) in the absence of treatment (**Fig 3D, dark blue and light green**). However, even in this scenario the number of current COVID-19 hospitalizations is expected to surpass 2000 (**Fig S4**) by Nov. 1 which corresponds to almost 40% occupancy. Inpatient treatment would lower deaths significantly but would provide no relief for the pressure on the health care system as the hospitalization rate would remain unchanged. Therefore, if 75% pC_PI is restored, adjunctive strategies are likely to be insufficient without reversing to greater restrictions of physical interactions.

### Adjunctive interventions could be effective if only 60% pre-COVID physical interactions are restored

A slowly escalating number of daily deaths is predicted in mid-September if test and isolate strategy (TI_S) is added to 60% pC_PI with ~35 deaths per day by November (**Fig 3C, light blue**). This strategy is predicted to reduce cumulative deaths projected by November 1 by 70% (BF) with median projected deaths (UI) across the set of parameterizations under all reopening plans remaining below 1000 (**Fig 4A**, light blue). More than 75% of the acceptable parameterizations implementing TI_S predict fewer than 10 deaths per day under the baseline reopening plan (**Fig 4B**, light blue) although in few simulations the maximum daily deaths go up to 30.

Comprehensive test and trace (TI_S+C) also allows gradual restoration of physical interactions to 60% of their pre-COVID levels for all age groups without significant increases in hospitalizations (**Fig S5**) or deaths (**Fig 3C and 4A**, light green). Under this scenario no more than 7 deaths (BF, UI range 2-8) should be expected daily (**Table S5**).

At 60% pC-PI, treatment of isolated cases (TIT_S) alone would lower the cumulative deaths by 81% (BF, range 0-92% UI) through November (**Fig 3C**, dark blue), reducing the maximum daily deaths to 13 (BF) with 90% of the acceptable parameterizations remaining below 10 deaths per day under the baseline reopening plan (**Table S5**). Adding treatment to isolated cases and contacts (TIT_S+C) would allow at least 60% pC-PI without an increase in mortality (**Fig 3E**), with 90% of the parameterizations (BF and UI) remaining below 8 deaths per day by November 1. Current COVID-19 hospitalizations are expected to remain significantly lower than state mandated goal of 10% occupancy (**Fig S5**, dark green).

Inpatient treatment is unlikely to prevent virus resurgence even at 60% pC-PI with 65 daily deaths (BF) expected at the peak by November 1 in the baseline scenario and the threshold of 15 deaths being reached by early October (UI range, mid-August to not reached by the end of December). Interval mass testing of the population at levels feasible to date (0.5% daily) with isolation of cases would have a limited impact on the trajectory of deaths relative to no intervention at 60% pC-PI (**Fig 3C**, pink).

### Feasibility of expanded testing strategies

A reasonable goal for any intervention is to keep the death rate below peak death rate in April (~15 / day) through November. This can be achieved even with a higher pC-PI provided that there is greater success of the test and isolate or test and trace strategies (**Fig 5**). We explored scenarios with different levels of random (mass) testing and targeted (symptomatic) testing and evaluated the percent of parameterizations (from 0 to 100%) for which daily deaths remain below 15 through Nov. 1. Our analysis suggests that for safe restoration of 60% pC_PI, at least 2.5% of the population would need to be tested daily (roughly 55, 000 tests per day) to keep daily deaths below 15 (i.e., 80% of parameterizations below threshold, **Fig 5A**). The testing proportion increases further if larger fraction of pC_PI is allowed during reopening, with not even 4.5% daily testing being sufficient at 70% pC_PI. At 70% pC_PI, diagnosing 20% of symptomatic cases daily will achieve a similar level of mortality reduction (i.e., 80% of parameterizations below threshold) without contact tracing (**Fig 5B**) as diagnosing only 10% of symptomatic cases daily if effective contact tracing is assumed (**Fig 5C**).

**Figure 5.**
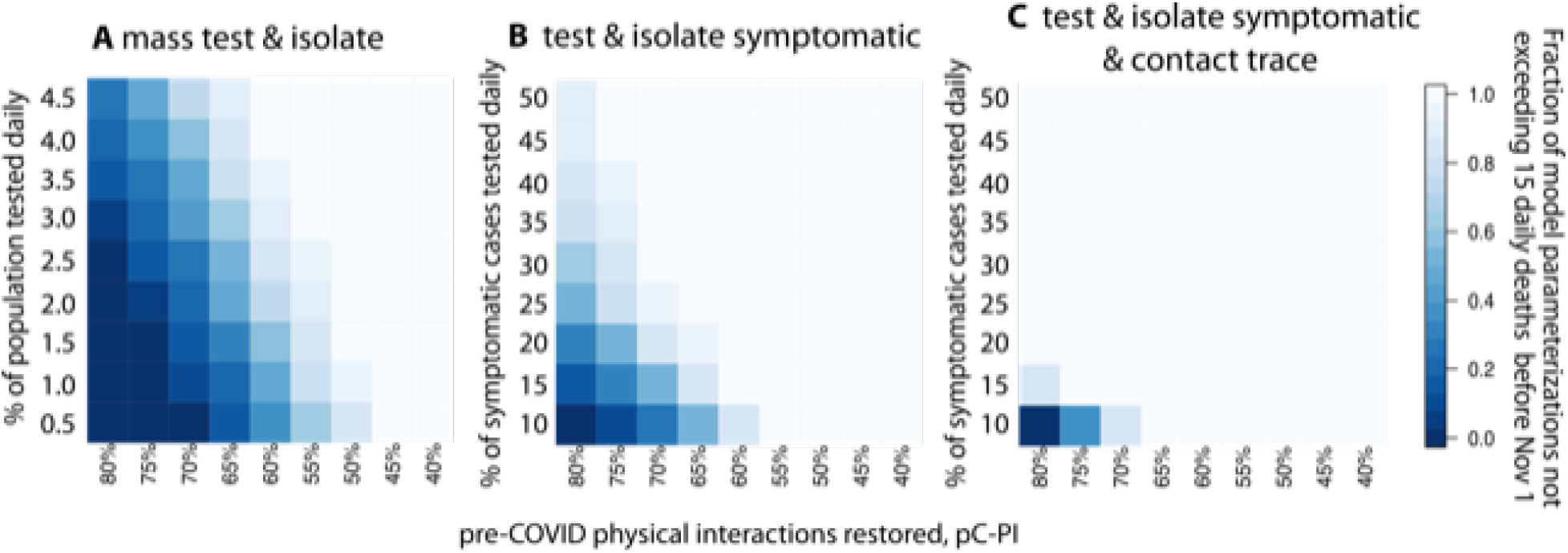
Model projections under different levels of diagnostic rates among. A) asymptomatic, pre-symptomatic and symptomatic cases due to mass testing programs; B)-C) symptomatic cases due to test & isolate programs. Reopening plan is implemented between May 15 and July 15 by restoring <u>different levels of pC PI</u> in all age groups on the x-axis. Heatmap represent the proportion of parameter sets (Ul)(from 0 to 1) for which daily deaths remain at or below 15 through November 1.

### Impact of isolating elderly populations

Our analysis suggests that protecting seniors above age of 70, by keeping them under the physical distancing achieved during the lockdown, would have only minimal positive impact on epidemic trajectories (**Fig 4**). For instance, the test and isolate systematic infections (TI_S) intervention decreases the cumulative death count by 12% (BF, range 1-14% UI) by Nov. 1 (**Fig 4A**) if seniors’ interactions are restricted compared to the baseline scenario, and maximum daily deaths decrease slightly (**Fig 4B**). Twelve percent of the parameterizations (UI) exceed 15 deaths daily compared to 17% in the baseline scenario. Unfortunately, the reduction of overall and maximum daily deaths is insufficient to make the disparate physical distancing strategies by age an effective policy in isolation.

### Impact of school reopening on the epidemic projections

Simulations of schools reopening reiterate further the need to maintain pC_PI below 60% and to include effective contact tracing to the intervention strategies against COVID-19. Our analysis demonstrates that opening schools is likely to more rapidly increase the death count to more than 15 per day if the only existing policy is diagnosing and isolating symptomatic cases (TI_S) with the BF parametrization crossing that threshold Sept. 22 and 46% of the parameterizations (UI) crossing that threshold compared to only 17% with the baseline scenario. In comparison, school reopening shows little impact if early infections are identified through contact tracing (TI_S+C) with only 1% of the parameterizations (UI) reaching 15 daily deaths by the end of 2020 (**Fig 4C**). This result suggests that at moderate pC_PI of 60%, school opening is only possible with successful contact tracing.

### Sensitivity analysis

We detected only slight sensitivity of cumulative deaths to the percentage of asymptomatic infections (parameter *p*) and the level of infectivity of those infections (parameter β_a_) after the end of the calibration period (**Fig 6A**) and also after the reopening plan is completed (**Fig 6B**). This suggests that our mortality projections are not influenced by the uncertainty in the asymptomatic assumptions. Similarly, the assumed asymptomatic infectivity does not impact the projected cumulative incidence over time (**Fig 6C, D**). Conversely, there is a clear association of increasing cumulative incidence with the increase of the fraction of asymptomatic infections. We estimate that 2.1% (median, range 1.0-10.2% UI) of the population is expected to be infected with SARS CoV-2 by Sept. 15 (assuming β_a_ = 1) if only 10% of the infections are asymptomatic compared to 2.7% (median, range 1.2-11.1% UI) and 5.0% (median, range 2.0-18.3% UI) if 20% and 50% of the infections are asymptomatic, respectively. The uncertainty ranges highlight the difficulty of determining the percentage of asymptomatic infections.

**Figure 6.**
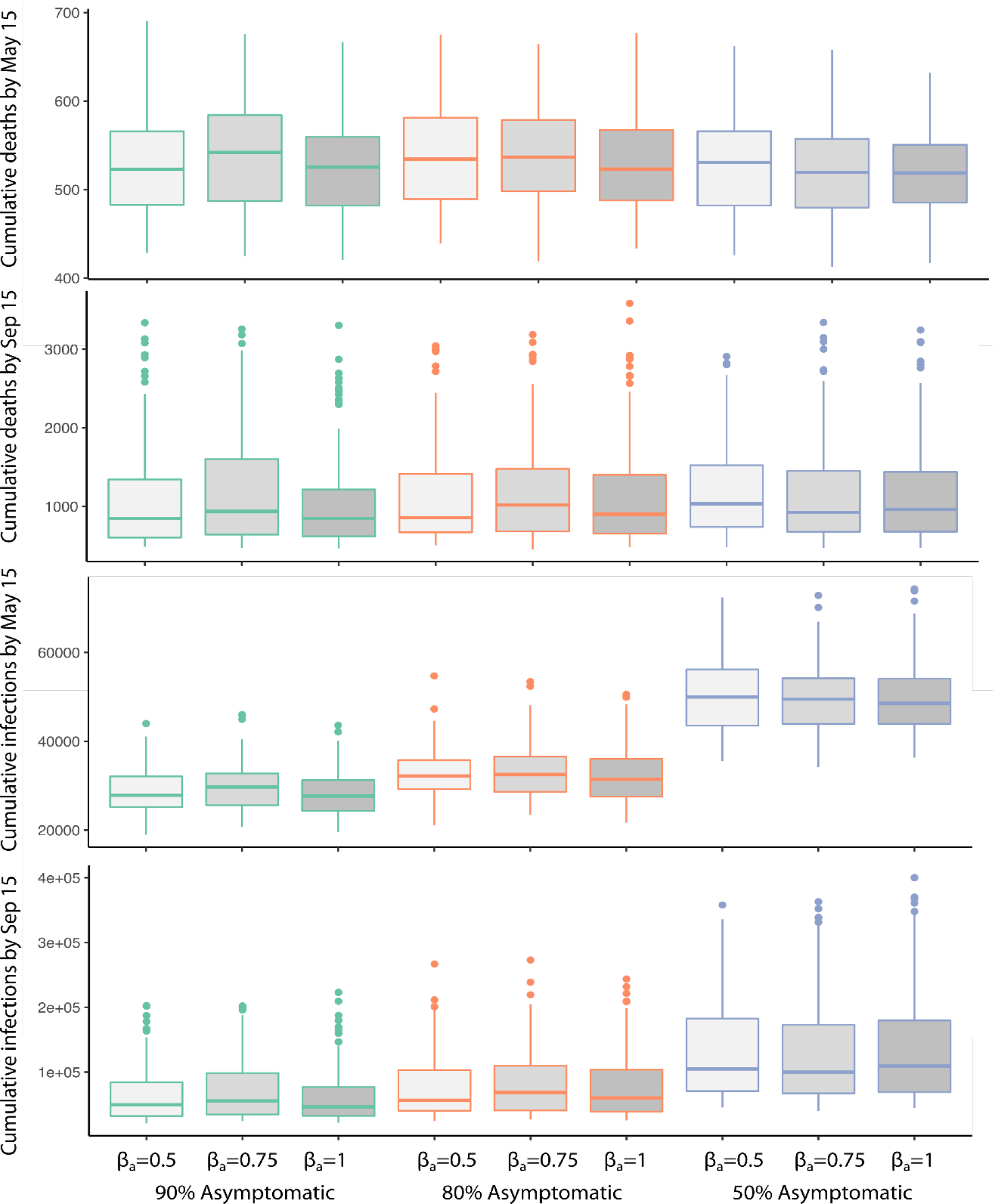
Model projections with different asymptomatic assumptions. A)-B) cumulative deaths by May 15 and Sept.15; C)-D) cumulative incidence at May 15 and Sept 15. Reopening plan is implemented between May 15 and July 15 by restoring <u>60% of pC PI</u> in all age groups on the x-axis. Boxplots show median and interquartile range (IQR) with whiskers extending to the smallest/largest value no further than 1.5 * IQR based on 100 accepted parameter sets per scenario.

## Discussion

Safely resuming economic and educational activities while avoiding a significant SARS-CoV-resurgence will require precise tuning the amount of physical interactions and will also be contingent upon adjunctive interventions such as improved testing and contact tracing. In this study, we used a mathematical model, specifically parameterized to King County, WA to assess the future trajectory of cases and deaths under a variety of reopening scenarios. We demonstrate that the prevention policies culminating with the “Stay Home, Stay Healthy” order were able to successfully reduce SARS-CoV-2 transmission by 65% and bring the effective reproductive number, which measures the average number of new infections transmitted from one infected individual at any specific time to a value below one, corresponding to a contracting epidemic. However, we predict that unless transmission is kept at or below 45% of pre-lockdown levels, the epidemic will continue its rapid rebound as has been observed in recent weeks. The increasing case count after some local and state restrictions were lifted combined with the lack of effective vaccine and other therapeutic products clearly suggest that it is necessary to prepare for a prolonged period of new sustainable endemic “normalcy”.

One of the most consistent findings from this modeling is that the effectiveness of individual interventions hinges on maintaining a certain degree of social distancing. Rapid testing and isolation alone may significantly curb transmission, but only if contacts are maintained at levels <60% pre-pandemic levels, or if testing is very widely implemented. We estimated that by May 15, more than 80% of symptomatically infected individuals remained undiagnosed. We demonstrate that close to 100% of symptomatically infected need to be tested and isolated to be possible to increase contacts to 70% of pre-pandemic levels without a surge in deaths.

We also investigated the ability of more comprehensive adjunctive interventions such as test and trace, post exposure prophylaxis and outpatient/inpatient treatments to containSARS-CoV-2 transmission in King County during and after the reopening period. Again, implementation of test and trace strategies is unlikely to prevent a surge in cases if 75% of the pre-COVID physical interactions are restored. However, this strategy would allow restoration to more than 60% of pre-COVID physical interactions without significant increases in daily cases and deaths, a scenario which may be feasible with stricter mask wearing policies.^25,26^

While outpatient therapies and post exposure prophylaxis measures are not yet licensed, these methods could further reduce the number of cases and deaths, and slightly increase the percentage of pre-pandemic contacts permitted before exponential growth in cases and deaths resume. Currently available inpatient treatments, such as remdesivir and dexamethasone are unlikely to control death because minimal SARS-CoV-2 transmission is generated by hospitalized patients.^19,27^ This is unlikely to change even as more potent and effective products become available.

As in other models^28^, we predict that mass testing interventions are also unlikely to be useful. We estimate that > 40, 000 daily tests would be needed, which is infeasible at the moment. This method is impractical mostly because of the low prevalence of the virus in any given time but also of poor detection of newly infected people who are not yet shedding virus and many new infections occurring between phased testing.

Another proposed strategy is to protect the highest risk members of society. However, we demonstrate that continuous restriction of seniors’ interactions at lockdown levels has a relatively small positive impact on the projected death rates. This finding serves as a warning that increased number of cases in younger demographics observed after reopening, puts the elderly at very high risk. Unfortunately, this risk has been realized in Florida and Arizona, where the prediction of our model of transmission diffusing into the older age groups has already occurred.

Reducing community transmission will be even more challenging if schools reopen in fall, potentially fueling new viral circulation. Therefore, we analyzed the rates of testing, case isolation and contact tracing which will allow for safe schools reopening in King County. Importantly, these rates are achievable only while the SARS-CoV-2 incidence remain low enough for the testing centers and contact tracers to be able to handle the demand in timely manner. Under certain scenarios including 60% pre-pandemic contact rate with active test and tracing school opening unfortunately tips the balance of transmission and causes death rates to increase dramatically. Together, our analysis argues for the urgent case reductions, high levels of physical distancing in the population at large, aggressive test and trace strategies, and careful evaluation of testing demand in order to make school openings safe enough to prevent new epidemic outbreaks.^29^

Our study has several limitations. As in most epidemics, SARS-CoV-2 transmission and acquisition risk is not distributed homogeneously. Individuals with more daily contacts, due to workplace or household size are at greater risk of infection. Those deemed to be essential workers who cannot physically distance will also be less responsive to state stay-at-home orders. Our model projections capture the heterogeneity in transmission by age but assume homogeneous risk of transmission and outcomes within age groups. Importantly for policy, this assumption means our current predictions are pessimistic. Another limitation of our analysis is that we apply uniform reduction of transmission across age groups due to social distancing. Contact rates of different age groups are surely affected differentially by the restrictions imposed to prevent the spread of SARS-CoV-2 (school closures, work-from-home policy, etc.). However, lack of local data prevented more detailed analysis. Implementing separate contact matrices for each transmission venue (home, school, work, etc.) in the future will allow more precision. Diagnostic rates are similarly simplified here. Some recent data suggest that time-varying age-specific diagnostic rate are more appropriate and will be included and informed by the number of tests and the fraction positive reported daily in the next model iteration.

Our analysis gives hope that widespread testing, case isolation and contact tracing may allow for coexistence with SARS-CoV-2 including safe schools reopening in King County without a virus resurgence which will overburden the health system and require further lockdowns. However, this will require continued physical distancing and a disciplined and coordinated effort to interrupt transmission prior to school opening followed by vigilant monitoring with broadly implemented contact tracing after the students return to school.

## Data Availability

The analysis was based on publicly available data.

